# Longitudinal Study after Sputnik V Vaccination Shows Durable SARS-CoV-2 Neutralizing Antibodies and Reduced Viral Variant Escape over Time

**DOI:** 10.1101/2021.08.22.21262186

**Authors:** María M. Gonzalez Lopez Ledesma, Lautaro Sanchez, Diego S. Ojeda, Santiago Oviedo Rouco, Andres H. Rossi, Augusto Varese, Ignacio Mazzitelli, Carla A. Pascuale, Esteban A. Miglietta, Pamela E. Rodríguez, Horacio M. Pallarés, Guadalupe S. Costa Navarro, Julio J. Caramelo, Paul W Rothlauf, Zhuoming Liu, Louis-Marie Bloyet, Marjorie Cornejo Pontelli, Natali B. Rasetto, Shirley D. Wenker, Lila Y. Ramis, Magalí G. Bialer, María Jose de Leone, C. Esteban Hernando, Luciana Bianchimano, Antonella Rios, María Soledad Treffinger Cienfuegos, Diana R. Rodriguez García, Yesica Longueira, Natalia Laufer, Diego Alvarez, Ana Ceballos, Valeria Ochoa, Cecilia Monzani, Gariela Turk, Melina Salvatori, Jorge Carradori, Katherine Prost, Alejandra Rima, Claudia Varela, Regina Ercole, Rosana I. Toro, Sebastian Gutierrez, Martín Zubieta, Dolores Acuña, Mercedes S. Nabaes Jodar, Carolina Torres, Laura Mojsiejczuk, Mariana Viegas, Pilar Velazquez, Clarisa Testa, Nicolas Kreplak, Marcelo Yanovsky, Sean Whelan, Jorge Geffner, Marina Pifano, Andrea V. Gamarnik

**Affiliations:** Fundación Instituto Leloir-CONICET, Av Patricias Argentinas 435, Buenos Aires, Argentina; Universidad de Buenos Aires, Facultad de Medicina, Instituto de Investigaciones Biomédicas en Retrovirus y SIDA (INBIRS-CONICET), Argentina; Department of Molecular Microbiology, Washington Univ. School of Medicine, St. Louis, USA; Biobanco de Enfermedades Infecciosas (INBIRS-UBA-CONICET), Argentina; Instituto de Investigaciones Biotecnológicas, UNSAM-CONICET, Argentina; Laboratorio Lemos S.R.L, Buenos Aires, Argentina; Hospital Interzonal General de Agudos Dr Pedro Fiorito, Provincia de Buenos Aires, Argentina; Hospital Interzonal General de Agudos Evita Provincia de Buenos Aires, Argentina; Hospital Interzonal General de Agudos Prof. Dr. Rodolfo Rossi, Prov. Buenos Aires, Argentina; Hospital Interzonal Especializado de Agudos y Crónicos San Juan de Dios, Provincia de Buenos Aires, Argentina; Hospital Interzonal General de Agudos San Roque, Provincia de Buenos Aires, Argentina; Hospital Interzonal General de Agudos San Martín, Provincia de Buenos Aires, Argentina; Hospital de Alta Complejidad El Cruce “Nestor Kirchner”, Provincia de Buenos Aires, Argentina; Hospital General de Niños Dr. Ricardo Gutierrez e Instituto de Investigaciones en Bacteriología y Virología Molecular, Fac de Farmacia y Bioquímica, UBA, Argentina; Ministerio de Salud de Provincia de Buenos Aires, Argentina

**Author notes:** Correspondence should be addressed to Andrea V. Gamarnik, Fundación Instituto Leloir, Buenos Aires, Argentina. These authors contributed equally to this work (María M. Gonzalez Lopez Ledesma, Lautaro Sanchez, Diego S. Ojeda, Santiago Oviedo Rouco).

## Abstract

Recent studies have shown a temporal increase in the neutralizing antibody potency and breadth to SARS-CoV-2 variants in coronavirus disease 2019 (COVID-19) convalescent individuals. Here, we examined longitudinal antibody responses and viral neutralizing capacity to the B.1 lineage virus (Wuhan related), to variants of concern (VOCs: Alpha, Beta, Gamma, and Delta) and a local variant of interest (VOI: Lambda) in volunteers receiving the Sputnik V vaccine in Argentina. A collection of 472 serum samples obtained between January and September 2021 was used. The analysis indicates that while anti-spike IgG levels significantly wane over time, the neutralizing capacity to the first-wave linages of SARS-CoV-2 and VOC are maintained within four months of vaccination. In addition, an improved antibody cross-neutralizing ability to circulating variants of concern (Beta, Gamma and Delta) was observed over time of vaccination. The viral variants that displayed higher escape to neutralizing antibodies with respect to the original virus (Beta and Gamma variants) were the ones showing the largest increase in susceptibility to neutralization over time after vaccination. Our observations indicate that serum neutralizing antibodies are maintained for at least four month and show a reduction of VOC escape over time of vaccination.

## INTRODUCTION

The coronavirus disease 2019 (COVID-19) pandemic is devastating economies and healthcare systems worldwide and has caused more than 4.4 million deaths by August 2021 (World Health Organization, 2021). Mass vaccination offers the possibility of halting this global burden. However, the limited vaccine supply and inequalities in vaccine accessibility create a need to increase international cooperation. An additional challenge in combatting COVID-19 has been the emergence in the late 2020 of new viral variants around the world with variable transmissibility, replication, and/or resistance to neutralizing antibodies. Viral variants harboring mutations in the spike protein may compromise vaccine immune control, and the rapid spread of these variants could undermine current efforts to end the pandemic. Thus, constant surveillance of viral variant emergence and documentation of immune responses elicited by different vaccine platforms are fundamental to optimizing pandemic control measures.

The humoral immune response elicited by SARS-CoV-2 infection and vaccination is a relevant marker of protection against subsequent viral encounters (Earle et al., 2021; Khoury et al., 2021; Krammer, 2021). Recent reports have provided important information regarding antibody durability and maturation processes in infected patients (Dispinseri et al., 2021; Gaebler et al., 2021; Wang et al., 2021). Antibody titers against SARS-CoV-2 were shown to wane over time for COVID-19 convalescent individuals, while antibody maturation increased the neutralization potency to the original SARS-CoV-2 and variants of concern (VOCs; Moriyama et al., 2021; Muecksch et al., 2021). This phenomenon has not been well described for vaccinated individuals. In this study, we evaluated the humoral response over time and the neutralizing potency of antibodies elicited by Sputnik V vaccination in Argentina. Sputnik V (Gam-COVID-Vac) is an heterologous recombinant adenovirus (rAd type 26 and rAd type 5) based vaccine (Logunov et al., 2020, 2021). Evaluation of the immune response over time up to four months after vaccination indicates that anti-spike antibody levels wane, but neutralization capacity is maintained, not only for the ancestral SARS-CoV-2 but also for widely and locally circulating viral variants. These data support the notion that antibody affinity maturation and limitation of VOC escape may occur over time after Sputnik V vaccination.

## RESULTS

We evaluated the longitudinal anti-spike IgG antibody level and viral neutralizing capacity to SARS-CoV-2 VOCs in 118 volunteers (see Supplementary Table 1) receiving the complete two-dose regimen of the Sputnik V vaccine in Argentina, as a continuation of our recent report (Rossi et al., 2021). A collection of 472 serum samples were initially obtained between January and September 2021. Plasma samples were taken at four time points: before vaccination (baseline), and at 21, 42 and 120 days after the initial vaccination (second dose was applied at 21 days). There was 100% volunteer adherence. The levels of IgG antibodies against the complete spike protein were measured by titration (Ojeda et al., 2021) and quantification was carried out using the WHO International Antibody Standard (Kristiansen et al., 2021a). According to the presence of antibodies at baseline, samples were divided in two groups, without (Group 1) or with (Group 2) previous SARS-CoV-2 infection. Virus neutralizing antibodies were evaluated using two systems: a pseudotyped VSV spike-expressing GFP (Case et al., 2020) for the Wuhan, Alpha (United Kingdom), Beta (South Africa), Gamma (Manaos), and Delta (India) variants; and the live SARS-CoV-2 using local isolates for the B.1 lineage (Wuhan related) and regional circulating SARS-CoV-2 variants (Alpha, Gamma, and Lambda).

### Sustained viral neutralization capacity over time upon Sputnik V vaccination

A longitudinal analysis of 118 volunteers vaccinated with the two-dose regimen of Sputnik V showed that IgG levels declined over a period of 4 months. The geometric mean (GM) of international units of IgG anti-spike antibodies per ml (IU/ml) for the group that was seronegative (naïve) at baseline (Group 1, *N* = 88) declined from 758 (CI95%, 574–1001) at 42 days to 228 (CI95%, 166–314) by 120 days after the initial vaccination (**Figure 1A**). IgG level waning was also observed in participants who were seropositive (due to prior infection) at baseline (Group 2, **Figure S1**). For this group, the GM of antibody (IU/ml) was the highest after the first dose of the vaccine, 9429 (CI95%, 6303– 14105), and declined to 5193 (CI95%, 3390–7960) and 2719 (CI95%, 1706–4333) at 42 and 120 days after the initial vaccination, respectively. The geometric mean half-maximal neutralizing titer (GMT IC50%) of Group 1 samples at 42 and 120 days after the initial vaccination was 116 (CI95%, 85–159) and 76 (CI95%, 51–114). No significant reduction in neutralizing capacity was observed over this period (**Figure 1B**).

**Figure 1.**
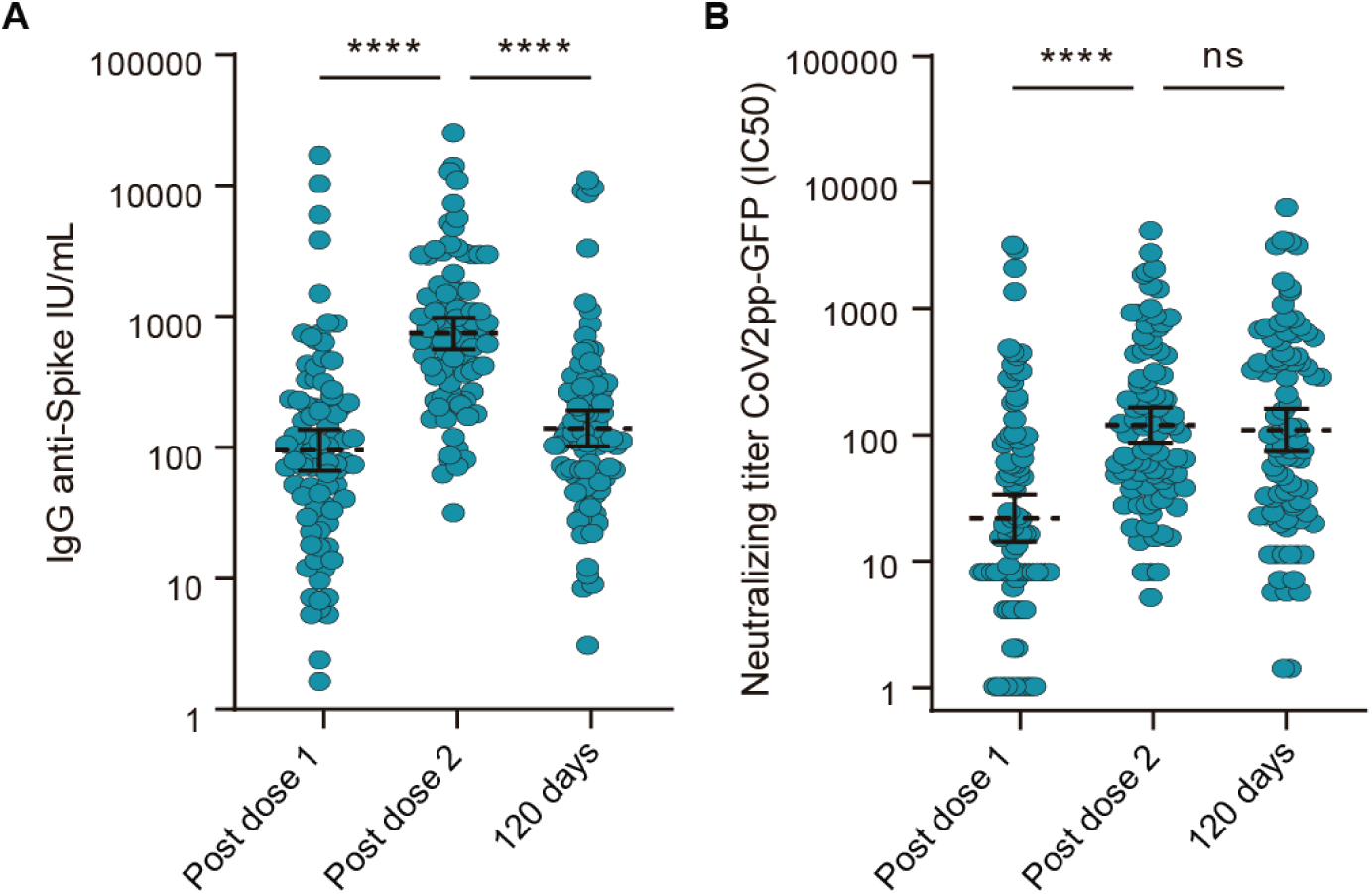
Immune response to the Sputnik V vaccine in naïve participants. (A) IgG anti-spike antibody levels quantified according to the WHO International Antibody Standard (*N* = 88). Antibodies were measured 21 (*N*=88), 42 (21 days post second dose) (*N*=88) and 120 (*N* = 88) days after the initial vaccination. (B) Neutralizing titers measured at 50% inhibition against the pseudotyped virus (CoV2pp GFP) for the same cohort as in A. The geometric means with 95% confidence intervals are shown. Wilcoxon matched paired test was used. Statistical significance is shown with the following notations: \*\*\*\**p* < 0.0001; \*\*\**p* < 0.001; ns, not significant.

The data show that, although the total amount of IgG anti-spike decreases as a function of time after Sputnik V vaccination, the neutralizing capacity in naïve individuals was maintained.

### Reduced VOC escape over time after Sputnik V vaccination

We then evaluated the serum neutralizing activity to circulating VOCs elicited by Sputnik V vaccination. Viral infection inhibition was assessed using two systems a VSV-based pseudotyped virus and the isolated SARS-CoV-2. The VSV-based system encoded GFP and was pseudotyped with the spike protein corresponding to the original SARS-CoV-2 (Wuhan), and the Alpha (lineage B.1.1.7), Beta (lineage B.1.351), Gamma (lineage P.1), and Delta (lineage B.1.617.2) variants, which were initially identified in United Kingdom, South Africa, Manaos, and India, respectively. Serum samples collected 42 days after vaccination showed a 2.5- and 5.1-fold decrease in neutralizing activity against the Alpha and Delta variants in comparison with the Wuhan related pseudotyped virus (*p* = 0.002 and *p* < 0,0001, respectively) (**Figure 2A**). The samples were less effective at neutralizing the Beta and Gamma variants (19.2- and 13.8-fold reduction in neutralizing activity, *p* < 0.0001 and *p* < 0.0001, respectively).

**Figure 2.**
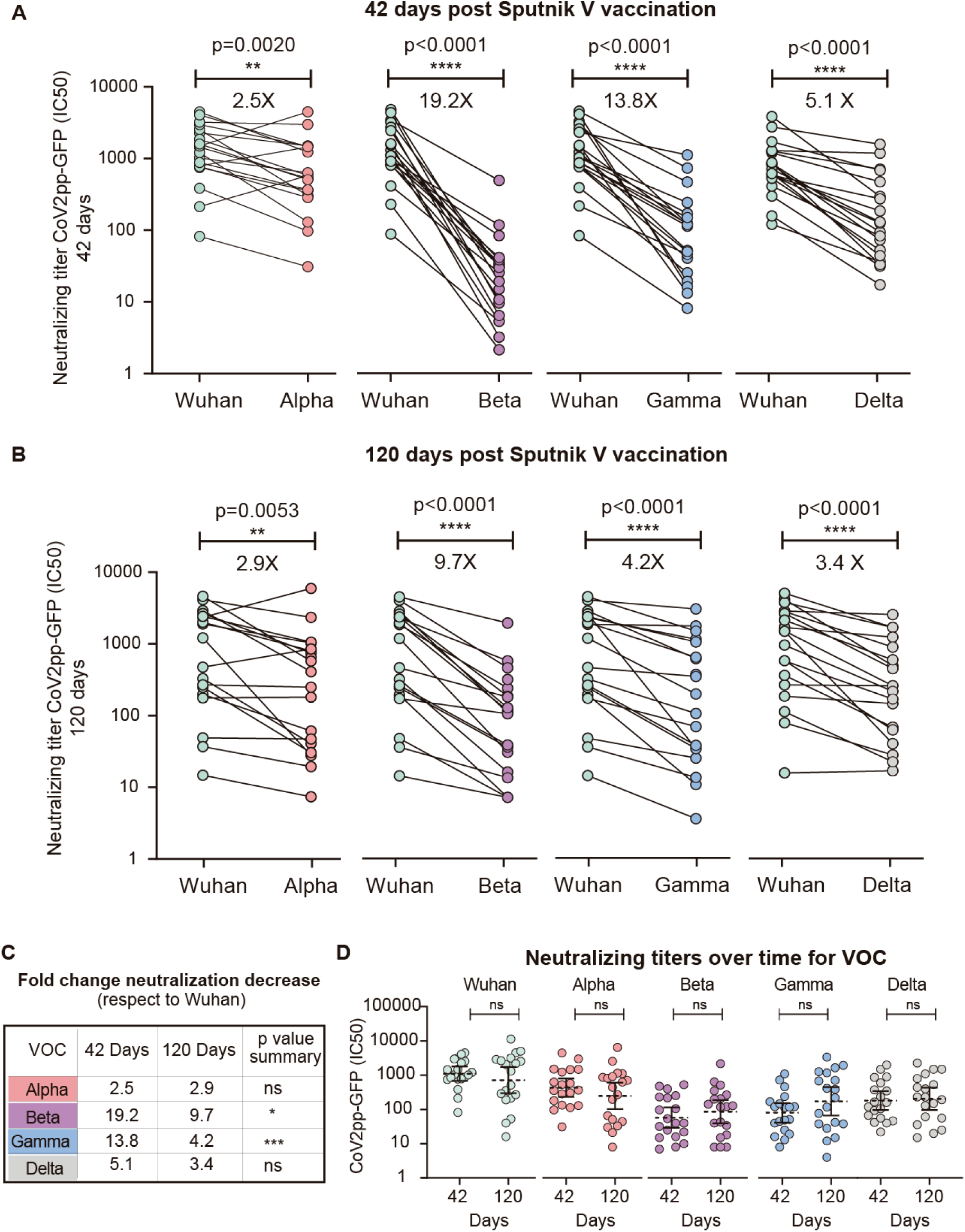
Longitudinal analysis of neutralizing capacities of 19 sera samples from Sputnik-vaccinated participants for each variant of concern (VOC). Half-maximal neutralizing titers (IC50%) against VOCs using pseudotyped viruses (CoV2pp GFP), shown as fold change reduction normalized to the IC50% titer against the original virus. Sera collected 42 (A) or 120 (B) days after the initial vaccination were used for neutralization assays with the Alpha, Beta, Gamma, and Delta variants, as indicated in each case. A Wilcoxon matched paired test was used to analyze the data shown in A and B. (C) Comparison of fold change neutralization titer decrease for each VOC respect to the Wuhan virus at 42 and 120 days after vaccination. The significance of the reduction is indicated on the right. (D) Neutralizing capacity at 42 and 120 days after initial vaccination for each variant as indicated. For non-paired samples analysis in panel C and D the Mann-Whitney U test was used. Statistical significance is shown with the following notations: ****, *p* < 0.0001; ***, *p* < 0.001; *, *p* < 0.05; ns, not significant.

Serum samples collected 120 days after vaccination showed a 2.9-, 9.7-, 4.2-, and 3.4-fold decrease in neutralizing activity against the Alpha, Beta, Gamma, and Delta variants, respectively, compared with the Wuhan virus (**Figure 2B**). Interestingly, a significant reduction of neutralization escape was observed over time (from 42 to 120 days after vaccination) for the Beta and Gamma VOCs (**Figure 2C**). In addition, the level of neutralizing capacities over time was maintained for all the variants tested (**Figure 2D**).

The Lambda variant was initially detected in late December 2020 in South America (Andina, lineage C.37)(Romero et al., 2021). This novel sublineage within B.1.1.1, with a convergent deletion in the ORF1a gene (Δ3675–3677) and a novel deletion in the spike gene (Δ246–252, G75V, T76I, L452Q, F490S, T859N), rapidly spread in the region, replacing the Alpha variant and reaching frequencies in Argentina as high as 48% (Proyecto País, 2021). The Lambda, Alpha, and Gamma variants are the main SARS-CoV-2 viruses currently circulating in Argentina. The antibody neutralizing activity elicited by the Sputnik V vaccine was analyzed using the original local isolate B.1 (Wuhan related) virus and local isolates of the viral variants. A subset of 40 randomly selected group 1 volunteers were used for this analysis. Neutralizing titers were defined as the highest serum dilution that failed to elicit a cytopathic effect on the cell monolayer (**Figure 3**).

**Figure 3.**
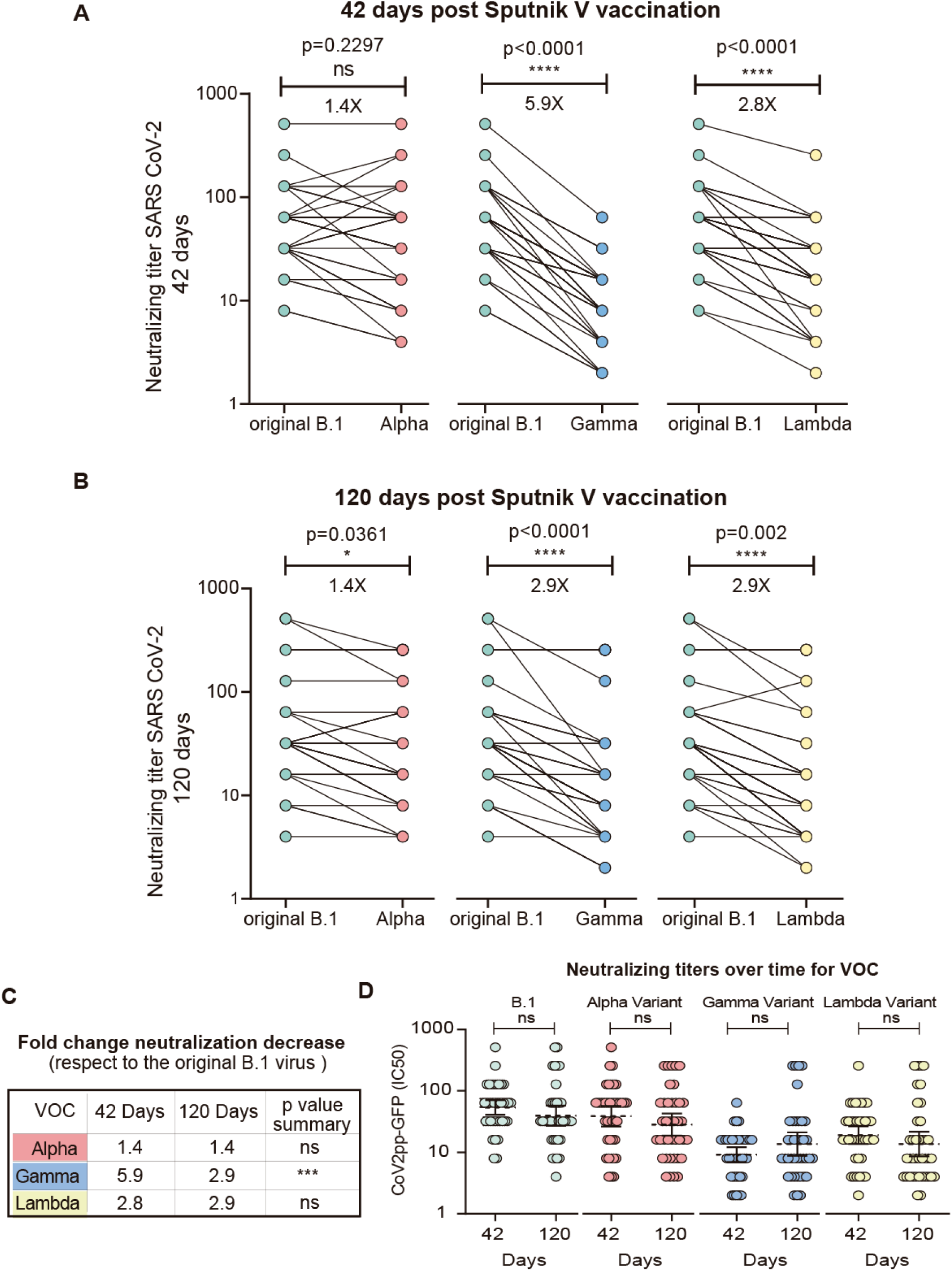
Longitudinal analysis of the neutralizing capacities of sera from Sputnik V-vaccinated individuals for each variant of concern (VOC) using viral isolates circulating in Argentina. The half-maximal neutralizing titer (IC50%) for VOCs using a replicating SARS-CoV-2 was assessed and shown as fold change reduction with respect to that for the original B.1 virus. Neutralizing Titers were defined as the highest serum dilution without any cytopathic effect on the monolayer. Sera (*N*=40) collected 42 days (A) or 120 days (B) after initial immunization were used for neutralization assays with Alpha, Gamma and Lambda variants, as indicated in each case. Wilcoxon matched paired test was used in panels A and B. (C) Comparison of fold change neutralization titer decrease for each VOC respect to the original B.1 virus at 42 and 120 days after vaccination. The significance of the reduction is indicated on the right. (D) Neutralizing capacity calculated at 42 and 120 days after initial immunization is shown for each variant as indicated. For non-paired samples analysis in panel C and D the Mann-Whitney U test was used. Statistical significance is shown with the following notations: ****, *p* < 0.0001; ***, *p* < 0.001; *, *p* < 0.05; ns, not significant.

The virus neutralizing capacity of serum from Sputnik V-vaccinated individuals was slightly lower for the Lambda variant than for the original B.1 virus (2.8- and 2.9-fold change, at 42 and 120 days after the first vaccination dose, respectively). For the Alpha variant, no significant (at 42 days after the first dose) or mild (at 120 days after the first dose) escape to neutralization was observed (**Figure 3A and B**). By contrast, sera from vaccinated volunteers were less effective against the Gamma variant, showing a 5.9- and 2.9-fold reduction of inhibition in samples collected at 42 and 120 days after the initial vaccination, respectively (**Figure 3C)**. Neutralization capacity was maintained over time after Sputnik V vaccination for all SARS-CoV-2 variants analyzed (**Figure 3D**). Together, these data suggest an improved cross-neutralization capacity, limiting VOC antibody escape over time after vaccination.

## CONCLUSION

Unraveling the long-term kinetics of antibodies to SARS-CoV-2 and VOCs in vaccinated individuals is important for understanding protective immunity against COVID-19 and for devising effective control measures. Here, we investigated the longitudinal humoral response in 118 volunteers vaccinated with Sputnik V. Our data indicate that anti-spike IgG antibodies wane over 4 months after vaccination without loss of serum neutralizing capacity against the original SARS-CoV-2 variant. The neutralizing capacity of the antibodies elicited by the vaccine was either slightly reduced or not reduced at all for the Alpha, Lambda, and Delta variants, while partial escape was observed for the Beta and Gamma variants. Antibodies elicited in individuals vaccinated with the Sputnik V vaccine also exhibited increased cross-neutralization capacity over time to VOCs. This might reflect the maturation of the antibody response induced by Sputnik V vaccination. Similar cross-neutralization increase to the original and VOC viruses was recently observed in longitudinal studies of individuals previously infected with SARS-CoV-2 (Moriyama et al., 2021) and of those harboring cloned neutralizing antibodies derived from convalescent donors (Gaebler et al., 2021; Muecksch et al., 2021). These studies suggest that declining antibody titers may not be indicative of declining protection. In summary, our studies support an increase of cross-neutralization to circulating SARS-CoV-2 variants, reducing the viral scape, in the months following Sputnik V vaccination. Further studies to evaluate efficacy over time for VOC in vaccinated individuals will be necessary to define correlates of protection with antibody and neutralization titers.

## METHODS

### Patient and sample origin

This study monitors the humoral immune response over time post immunization with Sputnik V vaccine in 118 health care workers from Buenos Aires province, Argentina. Patient information is given in Supplementary Table 1. Blood was collected by venipuncture into SST tubes (BD Sciences) for serum and stored at 20ºC.

### Ethics

Study enrollment started in January 2021 and is ongoing. Ethical approval was obtained from the central committee of the Ministry of Health of Buenos Aires and all participants provided written informed consent prior to collection of data and specimen (Cod#2021-00983502). All specimens were de-identified prior to processing and antibody testing for all serum specimens.

### Cohort description

The cohort were 31.4% male and 68.6% female with an average age of 49.3 years (range 22-76 years).Information about ethnicity was not collected.

Sequential serum samples were collected at five time points: before vaccination (baseline), and at 21, 42 and 120 days after the initial vaccination starting in January 2021. Volunteers were divided into two groups: without (Group 1) or with (Group 2) previous SARS-CoV-2 infection. Volunteers were classified according to the result of an ELISA IgG anti-Spike test at baseline.

### Cell lines

Vero-CCL81 cells (ATCC) and 293T ACE2/TMPRSS2 cells (kindly provided by Dr. Benhur Lee) were used. Cell were cultured at 37ºC in 5% CO2 in Dulbecco’s Modified Eagle’s high glucose medium (Thermo Fisher Scientific) supplemented with 10% fetal bovine serum (FBS)(GIBCO).

### Viruses

SARS-CoV-2 pseudotyped particles (CoV2pp-GFP) expressing spike protein were generated in Sean Whelan laboratory (Case et al., 2020). The S gene of SARS-CoV-2 isolate Wuhan-Hu-1 (GenBank MN908947.3) and recombinant VSVs expressing variants of the SARS-CoV-2 spike, including B.1.1.7 (GenBank OU117158.1), B.1.351 (GenBank MZ212516.1), and P.1 (GISAID EPI_ISL_804823) were generated see below (Case et al., 2020).

The B.1, Alpha (B.1.1.7), Gamma (P.1) and Lambda (C.37) isolates were obtained from nasopharyngeal isolates. All viruses were passaged once in Vero cells and subjected to Sanger sequencing to confirm the introduction and stability of substitutions. All virus experiments were performed in an approved biosafety level 3 facilities.

### Recombinant VSV

Briefly, recombinant VSVs were rescued by infecting BSRT7/5 cells with vaccinia virus vTF7-3 and subsequently transfecting them with T7-driven support plasmids encoding VSV N, P, L, G, and VSV genomic cDNAs. Supernatants were harvested 72 h post-infection, cellular debris was removed by centrifugation (5 min and 1,000 x g), and supernatants were passed through 0.22 μm filters. Supernatants were plaque-purified on Vero-CCL81 cells. Individual clones were grown on Vero-CCL81 cells to generate P1 stocks. Working stocks were generated on Vero-CCL81 cells at 34°C. Viral stocks (VSV-eGFP-SARS-CoV-2), generated in Sean Whelan laboratory, were amplified in our laboratory using 293T ACE2/TMPRSS2 cells at an MOI of 0.01 in Dulbecco’s Modified Eagle’s medium containing 2% FBS at 37ºC. Viral supernatants were harvested upon extensive cytopathic effect and GFP positive cells. The media was clarified by centrifugation at 1,000 x g for 5 min. Viral stocks were titrated by fluorescence forming units per milliliter (UFF/ml) in Vero cell line. Aliquots were maintained at −80ºC.

### SARS-CoV-2

SARS-CoV-2 ancestral reference strain 2019 (GISAID Accession ID: EPI_ISL_499083) B.1 was obtained from Dr. Sandra Gallegos (InViV working group). Alpha (GISAID Accession ID: EPI_ISL_2756558) and Gamma (GISAID Accession ID: EPI_ISL_2756556) variants were isolated at INBIRS from nasopharyngeal swabs. Lambda (hCoV-19/Argentina/PAIS-A0612/2021 GISAID Accession ID: EPI_ISL_3320903) variants was isolated at INBIRS from a sample of nasopharyngeal swab kindly transferred by Dr. Viegas and Proyecto PAIS. Virus was amplified in Vero E6 cells and each stock was fully sequenced. Work with SARS-CoV-2 was approved by the INBIRS Institutional Biosafety Committee at Biosafety level 3 with negative pressure.

### Sequencing of the S gene

Viral RNA was extracted from VSV-SARS-CoV-2 mutant viruses using TRIzol LS Reagent (Thermo Fisher Scientific), and S was amplified using M-MLV Reverse Transcriptase (Thermo Fisher Scientific). The mutations were identified by Sanger sequencing (Applied Biosystems).

### SARS-CoV-2 antibody ELISA

Antibodies to SARS-CoV-2 spike protein were detected using an established two step ELISA previously described (Ojeda et al., 2021). This assay has plates coated with a mixture of spike and the receptor binding domain (RBD). The SARS-CoV-2 antibody concentration of each sample expressed in International Units/mL (UI/mL)(Kristiansen et al., 2021b) was calculated by extrapolation of the OD 450 nm value on a calibration curve. For construction of the calibration curve, we determined OD 450 nm of serial dilutions of the WHO International Standard for anti-SARS-CoV-2 immunoglobulin. The linear range used was 0.2-1.5 OD 450nm. Therefore, we performed serial dilutions of the samples in order to find conditions where the OD 450nm of each sample fit adequately in the linear range.

### SARS-CoV-2 spike pseudotyped VSV neutralization assay

To compare the neutralizing activity of volunteer’s sera against coronaviruses, neutralization assays were carried out with SARS-CoV-2 pseudotyped particles (CoV2pp-GFP), generated in Sean Whelan laboratory (Case et al., 2020). CoV2pp-GFP carries vesicular stomatitis virus as viral backbone, bearing the E gene in place of its G glycoprotein (VSV-eGFPSARS-CoV-2), and expresses full length wild-type or mutant spike variant on its envelope. Vero cells were used for these assays. Cells were maintained with DMEM high glucose with 10% FBS and were seeded in a 96-well plate the day before infection. Patient sera were heat inactivated at 56ºC for 30 minutes and serially diluted in DMEM high glucose medium. Serum neutralizations were performed by first diluting the inactivated sample 2-folds and continuing with a 2-fold serial dilution. A pre-titrated amount of pseudotyped particles was incubated with a 2-fold serial dilution of patient sera for 1 h at 37ºC prior to infection. Subsequently, cells were fixed in 4% formaldehyde containing 2 mg/mL DAPI nuclear stain (Invitrogen) for 1 hour at room temperature, and fixative was replaced with PBS. Images were acquired with the InCell 2000 Analyzer (GE Healthcare) automated microscope in both the DAPI and FITC channels to visualize nuclei and infected cells (i.e., eGFP-positive cells), respectively (4X objective, 4 fields per well, covering the entire well). Images were analyzed using the Multi Target Analysis Module of the InCell Analyzer 2000 Workstation Software (GE Healthcare). GFP-positive cells were identified in the FITC channel using the top-hat segmentation method and subsequently counted within the InCell Workstation software. Absolute inhibitory concentrations (absIC) values were calculated for all patient sera samples by modeling a 4-parameter logistic (4PL) regression with GraphPad Prism 8. The 4PL model describes the sigmoid-shaped response pattern. For clarity, it is assumed that the response can be expressed so that the slope increases as the concentration increase. Absolute inhibitory concentration (absIC) was calculated as the corresponding point between the 0% and 100% assay controls. Fifty % inhibition was defined by the controls for all the samples on the same plate. For example, the absIC50 would be the point at which the curve matches inhibition equal to exactly 50% of the 100% assay control relative to the assay minimum. A 4-parameter logistic (4PL) regression with GraphPad Prism 8 was used.

### Authentic SARS-CoV-2 neutralization assay

Serum samples were heat-inactivated at 56ºC for 30 min and serial dilutions from 1/2 to 1/8192 were incubated for 1hs at 37ºC in the presence of ancestral or variants of SARS-CoV-2 in DMEM 2% FBS. Fifty ml of the mixtures were then deposited over Vero cells monolayers for an hour at 37ºC (MOI = 0.01). Infectious media was removed and replaced for DMEM 2% FBS. After 72 h, cells were fixed with PFA 4% (4ºC 20min) and stained with crystal violet solution in methanol. The cytopathic effect (CPE) of the virus on the cell monolayer was assessed visually, if even a minor damage to the monolayer (1-2 « plaques») was observed in the well, this well was considered as a well with a manifestation of CPE. Neutralization titer was defined as the highest serum dilution without any CPE in two of three replicable wells.

### Quantification and statistical analysis

Antibody concentration, neutralizing titer and neutralizing potency index from volunteers of the same group were analyzed collectively. Neutralization assays were performed in biological duplicates

All statistical tests and plots were performed using GraphPad Prism 8.0 software. Comparison on non-paired determinations of antibody concentration and neutralizing titer made using two-tailed Mann Whitney U test in Figures 2C, 2D, 3C and 3D. Comparison of antibody concentration, and neutralizing titer were made using two-tailed Wilcoxon matched paired test in Figures 1A, 1B, 2A, 2B, 3A and 3B and S1. Statistical significance is shown in the figure legends with the following notations: ****,p < 0.0001; ***,p < 0.001; **,p < 0.01; *,p < 0.05; ns, not significant. Geometric means with 95% confidence intervals were calculated in Figures 1A, 1B, 2A, 2B, 3A, 3B and S1.

## Data Availability

Data will be available.

## Data availability statement

The datasets generated during and/or analyzed during the current study are available in the Mendeley Data repository, DOI:10.17632/v2ksr58dcv.1.

**Supplementary Table 1:** Patient information. Age range, gender and group classification.

| ID | Sex | Age range | Group clasification |
| --- | --- | --- | --- |
| 1 | F | 76-80 | 1 |
| 2 | M | 71-75 | 1 |
| 3 | M | 71-75 | 1 |
| 4 | M | 66-70 | 1 |
| 5 | M | 66-70 | 1 |
| 6 | F | 66-70 | 1 |
| 7 | M | 61-65 | 1 |
| 8 | M | 61-65 | 1 |
| 9 | M | 66-70 | 1 |
| 10 | F | 61-65 | 1 |
| 11 | F | 61-65 | 1 |
| 12 | F | 61-65 | 1 |
| 13 | M | 61-65 | 1 |
| 14 | M | 61-65 | 1 |
| 15 | F | 61-65 | 1 |
| 16 | M | 61-65 | 1 |
| 17 | F | 61-65 | 1 |
| 18 | M | 61-65 | 1 |
| 19 | F | 61-65 | 1 |
| 20 | F | 61-65 | 1 |
| 21 | F | 61-65 | 1 |
| 22 | M | 61-65 | 1 |
| 23 | M | 61-65 | 1 |
| 24 | F | 56-60 | 1 |
| 25 | F | 61-65 | 1 |
| 26 | M | 56-60 | 1 |
| 27 | F | 56-60 | 1 |
| 28 | F | 56-60 | 1 |
| 29 | F | 56-60 | 1 |
| 30 | F | 56-60 | 1 |
| 31 | M | 56-60 | 1 |
| 32 | F | 56-60 | 1 |
| 33 | F | 51-55 | 1 |
| 34 | F | 51-55 | 1 |
| 35 | M | 51-55 | 1 |
| 36 | M | 51-55 | 1 |
| 37 | M | 51-55 | 1 |
| 38 | M | 51-55 | 1 |
| 39 | F | 46-50 | 1 |
| 40 | F | 51-55 | 1 |
| 41 | M | 51-55 | 1 |
| 42 | F | 51-55 | 1 |
| 43 | F | 51-55 | 1 |
| 44 | F | 51-55 | 1 |
| 45 | M | 51-55 | 1 |
| 46 | M | 46-50 | 1 |
| 47 | F | 46-50 | 1 |
| 48 | F | 46-50 | 1 |
| 49 | F | 46-50 | 1 |
| 50 | F | 46-50 | 1 |
| 51 | M | 46-50 | 1 |
| 52 | F | 46-50 | 1 |
| 53 | F | 46-50 | 1 |
| 54 | F | 46-50 | 1 |
| 55 | F | 46-50 | 1 |
| 56 | F | 41-45 | 1 |
| 57 | F | 41-45 | 1 |
| 58 | F | 41-45 | 1 |
| 59 | M | 41-45 | 1 |
| 60 | F | 41-45 | 1 |
| 61 | F | 41-45 | 1 |
| 62 | F | 41-45 | 1 |
| 63 | F | 41-45 | 1 |
| 64 | F | 41-45 | 1 |
| 65 | M | 41-45 | 1 |
| 66 | F | 36-40 | 1 |
| 67 | F | 36-40 | 1 |
| 68 | M | 36-40 | 1 |
| 69 | F | 36-40 | 1 |
| 70 | M | 36-40 | 1 |
| 71 | F | 36-40 | 1 |
| 72 | F | 36-40 | 1 |
| 73 | M | 36-40 | 1 |
| 74 | F | 36-40 | 1 |
| 75 | F | 31-35 | 1 |
| 76 | M | 31-35 | 1 |
| 77 | M | 31-35 | 1 |
| 78 | F | 31-35 | 1 |
| 79 | F | 31-35 | 1 |
| 80 | F | 31-35 | 1 |
| 81 | F | 31-35 | 1 |
| 82 | F | 26-30 | 1 |
| 83 | F | 26-30 | 1 |
| 84 | F | 26-30 | 1 |
| 85 | F | 26-30 | 1 |
| 86 | F | 26-30 | 1 |
| 87 | F | 26-30 | 1 |
| 88 | F | 21-25 | 1 |
| 89 | M | 56-60 | 2 |
| 90 | M | 26-30 | 2 |
| 91 | F | 41-45 | 2 |
| 92 | F | 61-65 | 2 |
| 93 | F | 41-45 | 2 |
| 94 | M | 56-60 | 2 |
| 95 | F | 36-40 | 2 |
| 96 | M | 61-65 | 2 |
| 97 | M | 21-25 | 2 |
| 98 | F | 46-50 | 2 |
| 99 | F | 26-30 | 2 |
| 100 | F | 46-50 | 2 |
| 101 | F | 46-50 | 2 |
| 102 | M | 46-50 | 2 |
| 103 | F | 51-55 | 2 |
| 104 | F | 26-30 | 2 |
| 105 | F | 56-60 | 2 |
| 106 | F | 41-45 | 2 |
| 107 | F | 46-50 | 2 |
| 108 | F | 36-40 | 2 |
| 109 | F | 46-50 | 2 |
| 110 | F | 36-40 | 2 |
| 111 | F | 61-65 | 2 |
| 112 | F | 51-55 | 2 |
| 113 | F | 56-60 | 2 |
| 114 | F | 66-70 | 2 |
| 115 | F | 26-30 | 2 |
| 116 | F | 56-60 | 2 |
| 117 | M | 56-60 | 2 |
| 118 | F | 51-55 | 2 |

**Figure S1.**
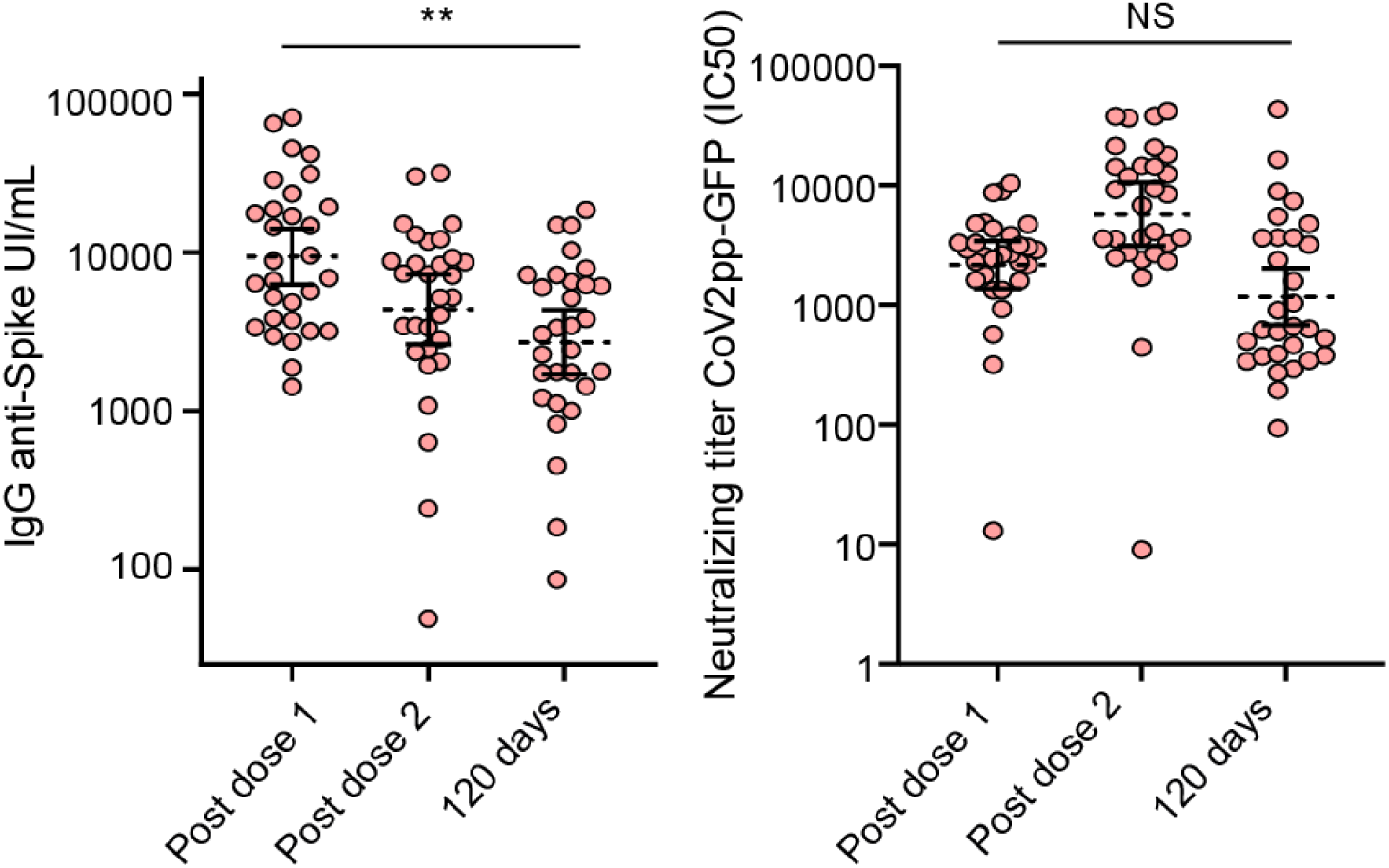
Immune response to the Sputnik V vaccine of seropositive participants at baseline. (A) IgG anti-SARS-CoV-2 spike levels quantified according to the WHO International Standard (*N* = 30). Antibodies were measured at 21, 42, and 120 days post the first dose of vaccine (post dose 1=21 days and post dose 2=42 days). (B) Neutralizing titers measured at 50% inhibition for the pseudotyped virus (CoV2pp GFP). Measurements were made 21, 42, and 120 days after first dose. Data were analyzed using a Wilcoxon matched paired test. Statistical significance is shown with the following notations: **, *p* < 0.01; NS, not significant.

## Acknowledgments

Authors are grateful to Dr Ana Fernandez Sesma and Dr Daniela Hozbor for helpful discussions and Dr Benhur Lee for providing the ACE2/TMPRSS2 expressing 293T cells. This work was supported by NIH (NIAID) R01.AI095175 and PICT-2017-1717, PICT-2015-2555 and Fondo para la Convergencia Estructural del Mercosur (FOCEM) to AVG and by the National Ministry of Science Technology and Innovation or Argentina. We are grateful to the following institutions and staff who contributed in sample collection: ***Biobanco de Enfermedades Infecciosas*:** Alejandro Czernikier, Yanina Ghiglione, Denise Giannone, María L. Polo, Florencia Quiroga, Belen Vecchione. We are thankful to Dr Guillermo Docena and members from the following hospitals: **HIEAC “San Juan de Dios”:** Andrea Gatelli, Sofia Di Bella, Agustina Martinez, Martina Ferioli, Francisco Echeverria, Ramiro Agüero, Ana Caproli, Karina Gil, Maria Colmeiro, Javier Rojas, Andres Angeletti, Luciano Malaissi, Edgardo Falcipieri, Julio Sachuen Leon; **HIGA “Dr. Rodolfo Rossi”:** Claudia Varela, Ángeles Baridon, Soledad Ocampo, Emanuel Zapata, Melina Cancela y Verónica Forneris. **HIGA “San Martín”:** Susana Marchetti, María Maxwell, Rosario Marcó, Cecilia Zolorzano, Micaela Nieva, Claudia Conta. **HIGA “Evita**”: Silvina Olivera, Isabel Desimone, Alejandra Musto. **HIGA “Dr. Pedro Fiorito”:** Aime Balanzino, Katherina Prost, Miriam Pereiro, Eliana Correa, Noelia Portillo, Cynthia Leguizamon, Alicia Quetglas. **HIGA “San Roque**”: Lucia Piumetti Agustina Venturi Grossi, Paula Gelpi, Anabella Masci, Sofía Padín, Jesica Tinto. **HAC “El Cruce-Néstor Kirchner”**: Mabel Skrypnik, Blanca Guevara, Virginia Aniasi, Alan Estigarribia

## REFERENCES

Case, J.B., Rothlauf, P.W., Chen, R.E., Liu, Z., Zhao, H., Kim, A.S., Bloyet, L.M., Zeng, Q., Tahan, S., Droit, L., et al. (2020). Neutralizing Antibody and Soluble ACE2 Inhibition of a Replication-Competent VSV-SARS-CoV-2 and a Clinical Isolate of SARS-CoV-2. Cell Host Microbe 28, 475-485.e5.

Dispinseri, S., Secchi, M., Pirillo, M.F., Tolazzi, M., Borghi, M., Brigatti, C., Angelis, M.L. De, Baratella, M., Bazzigaluppi, E., Venturi, G., et al. (2021). Neutralizing antibody responses to SARS-CoV-2 in symptomatic COVID-19 is persistent and critical for survival. Nat. Commun. 12, 1–12.

Earle, K.A., Ambrosino, D.M., Fiore-Gartland, A., Goldblatt, D., Gilbert, P.B., Siber, G.R., Dull, P., and Plotkin, S.A. (2021). Evidence for antibody as a protective correlate for COVID-19 vaccines. Vaccine 39, 4423–4428.

Gaebler, C., Wang, Z., Lorenzi, J.C.C., Muecksch, F., Finkin, S., Tokuyama, M., Cho, A., Jankovic, M., Schaefer-Babajew, D., Oliveira, T.Y., et al. (2021). Evolution of antibody immunity to SARS-CoV-2. Nature 591, 639–644.

Khoury, D.S., Cromer, D., Reynaldi, A., Schlub, T.E., Wheatley, A.K., Juno, J.A., Subbarao, K., Kent, S.J., Triccas, J.A., and Davenport, M.P. (2021). Neutralizing antibody levels are highly predictive of immune protection from symptomatic SARS-CoV-2 infection. Nat. Med. 27, 1205–1211.

Krammer, F. (2021). A correlate of protection for SARS-CoV-2 vaccines is urgently needed. Nat. Med. 27, 1147–1148.

Kristiansen, P.A., Page, M., Bernasconi, V., Mattiuzzo, G., Dull, P., Makar, K., Plotkin, S., and Knezevic, I. (2021a). WHO International Standard for anti-SARS-CoV-2 immunoglobulin. Lancet 397, 1347–1348.

Kristiansen, P.A., Page, M., Bernasconi, V., Mattiuzzo, G., Dull, P., Makar, K., Plotkin, S., and Knezevic, I. (2021b). WHO International Standard for anti-SARS-CoV-2 immunoglobulin. Lancet 397, 1347–1348.

Logunov, D.Y., Dolzhikova, I. V., Zubkova, O. V., Tukhvatullin, A.I., Shcheblyakov, D. V., Dzharullaeva, A.S., Grousova, D.M., Erokhova, A.S., Kovyrshina, A. V., Botikov, A.G., et al. (2020). Safety and immunogenicity of an rAd26 and rAd5 vector-based heterologous prime-boost COVID-19 vaccine in two formulations: two open, non-randomised phase 1/2 studies from Russia. Lancet 396, 887–897.

Logunov, D.Y., Dolzhikova, I. V, Shcheblyakov, D. V, Tukhvatulin, A.I., Zubkova, O. V, Dzharullaeva, A.S., Kovyrshina, A. V, Lubenets, N.L., Grousova, D.M., Erokhova, A.S., et al. (2021). Safety and efficacy of an rAd26 and rAd5 vector-based heterologous prime-boost COVID-19 vaccine: an interim analysis of a randomised controlled phase 3 trial in Russia. Lancet 397, 671–681.

Moriyama, S., Adachi, Y., Sato, T., Tonouchi, K., Sun, L., Fukushi, S., Yamada, S., Kinoshita, H., Nojima, K., Kanno, T., et al. (2021). Temporal maturation of neutralizing antibodies in COVID-19 convalescent individuals improves potency and breadth to circulating SARS-CoV-2 variants. Immunity 54, 1–12.

Muecksch, F., Weisblum, Y., Barnes, C.O., Schmidt, F., Schaefer-Babajew, D., Wang, Z., Lorenzi, J.C.C., Flyak, A.I., DeLaitsch, A.T., Huey-Tubman, K.E., et al. (2021). Affinity maturation of SARS-CoV-2 neutralizing antibodies confers potency, breadth, and resilience to viral escape mutations. Immunity 0, 1–16.

Ojeda, D.S., Gonzalez Lopez Ledesma, M.M., Pallarés, H.M., Costa Navarro, G.S., Sanchez, L., Perazzi, B., Villordo, S.M., Alvarez, D.E., Echavarria, M., Oguntuyo, K.Y., et al. (2021). Emergency response for evaluating SARS-CoV-2 immune status, seroprevalence and convalescent plasma in Argentina. PLOS Pathog. 17, e1009161.

Proyecto País (2021). Reporte N ° 15 : Vigilancia activa de variantes de SARS-CoV-2 en la CABA, provincias de Buenos Aires, Chaco, Córdoba, La Pampa, Neuquén y Santa Fe. Actualización al 13/07/2021. 2020, 1–11.

Romero, P.E., Dávila-Boarclay, A., Gonzáles, L., Salvatierra, G., Cuicapuza, D., Solis, L., Marcos, P., Huancachoque, J., Carhuaricra, D., Rosadio, R., et al. (2021). C.37: Novel lineage expanding in Peru and Chile, with a convergent deletion in the ORF1a gene (Δ3675-3677) and a novel deletion in the Spike gene (Δ246-252, G75V, T76I, L452Q, F490S, T859N).

Rossi, A.H., Ojeda, D.S., Varese, A., Sanchez, L., Gonzalez Lopez Ledesma, M.M., Mazzitelli, I., Juliá, A.A., Rouco, S.O., Pallarés, H.M., Costa Navarro, G.S., et al. (2021). Sputnik V Vaccine Elicits Seroconversion and Neutralizing Capacity to SARS CoV-2 after a Single Dose. Cell Reports Med. 2, 100359.

Wang, Z., Muecksch, F., Schaefer-Babajew, D., Finkin, S., Viant, C., Gaebler, C., Hoffmann, H.-H., Barnes, C.O., Cipolla, M., Ramos, V., et al. (2021). Naturally enhanced neutralizing breadth against SARS-CoV-2 one year after infection. Nat. 2021 5957867 595, 426–431.

World Health Organization (2021). WHO Coronavirus (COVID-19) Dashboard | WHO Coronavirus (COVID-19) Dashboard With Vaccination Data.

